# Suboptimal response to COVID-19 mRNA vaccines in hematologic malignancies patients

**DOI:** 10.1101/2021.04.06.21254949

**Authors:** Mounzer Agha, Maggie Blake, Charles Chilleo, Alan Wells, Ghady Haidar

## Abstract

Studies describing SARS-CoV-2 immune responses following mRNA vaccination in hematology malignancy (HM) patients are virtually non-existent. We measured SARS-CoV-2 IgG production in 67 HM patients who received 2 mRNA vaccine doses. We found that 46% of HM patients did not produce antibodies and were therefore vaccine non-responders. Patients with B-cell CLL were at a particularly high risk, as only 23% had detectable antibodies despite the fact that nearly 70% of these patients were not undergoing cancer therapy. HM patients should be counseled about the ongoing risk of COVID-19 despite vaccination. Routine measurement of post-vaccine antibodies in HM patients should be considered. Novel strategies are needed to prevent COVID-19 in these individuals.

---

Patients with hematologic malignancies are at high risk for coronavirus disease 2019 (COVID-19)-related complications, with mortality rates exceeding 30%^1-3^. These patients have also been shown to develop prolonged shedding of infectious severe acute respiratory syndrome coronavirus-2 (SARS-CoV-2), often lasting several months, and have been implicated in being sources of novel SARS-CoV-2 variants^4-7^. Such patients should be therefore be prioritized for primary prevention of COVID-19 via vaccination^8^. However, the performance of COVID-19 mRNA vaccines in hematological malignancy patients is unknown, as these individuals were excluded from COVID-19 vaccine clinical trials^9,10^.

To address these knowledge gaps, we measured SARS-CoV-2 antibody responses in patients with hematological malignancies seen at UPMC Hillman Cancer Center who have received two doses of either the mRNA-1273 (Moderna) or the BNT162b2 (Pfizer) vaccine. Patients with prior COVID-19 were excluded. Antibody assays were performed at the UPMC clinical laboratories using the semi-quantitative Beckman Coulter SARS-CoV-2 platform, which detects IgG against the Spike protein receptor-binding domain (RBD). These results are expressed as extinction coefficient (signal/cutoff) ratios and are interpreted as positive (≥ 1.00), equivocal (> 0.80 to < 1.00), or non-reactive (≤ 0.80)^11^. Reactive results are confirmed by the Siemens SARS-CoV-2 Total Ig Assay, which detects both IgM and IgG antibodies against RBD of the S1 subunit of the Spike protein^12^. For analysis, reactive results were defined as positive, and equivocal or non-reactive results were defined as negative. We calculated the proportion of patients with a positive versus negative result (vaccine responders versus non-responders, respectively) with 95% Coppler-Pearson exact confidence intervals and used χ^2^ or Wilcoxon Rank Sum testing for comparisons as appropriate. Analyses were performed using Stata version 16.1 (StataCorp) and GraphPad Prism 8.3.1. Institutional Board Review Approval was obtained.

Sixty-seven patients were included. Median age was 71 (interquartile range (IQR) 65 -77), and 47.8% (32/67) percent were female. Underlying malignancies were B-cell chronic lymphocytic leukemia (CLL, 19.4% (13/67)), lymphomas (31.3%, 21/67), multiple myeloma (43.3%, 29/67), and other myeloid malignancies (5.97%, 4/68) (**Table 1**). Thirty patients (44.8%) were undergoing therapy for their cancers, whereas 37 (55.2%) were under observation. Among the 62 patients whose vaccine type was available, 50.8% (34/67) and 41.8% (28/67) had received the BNT162b2 or mRNA-1273 vaccines, respectively. Median duration from the 2^nd^ vaccine dose to the antibody test was 23 days (IQR 16 -31 days).

**Table 1.**
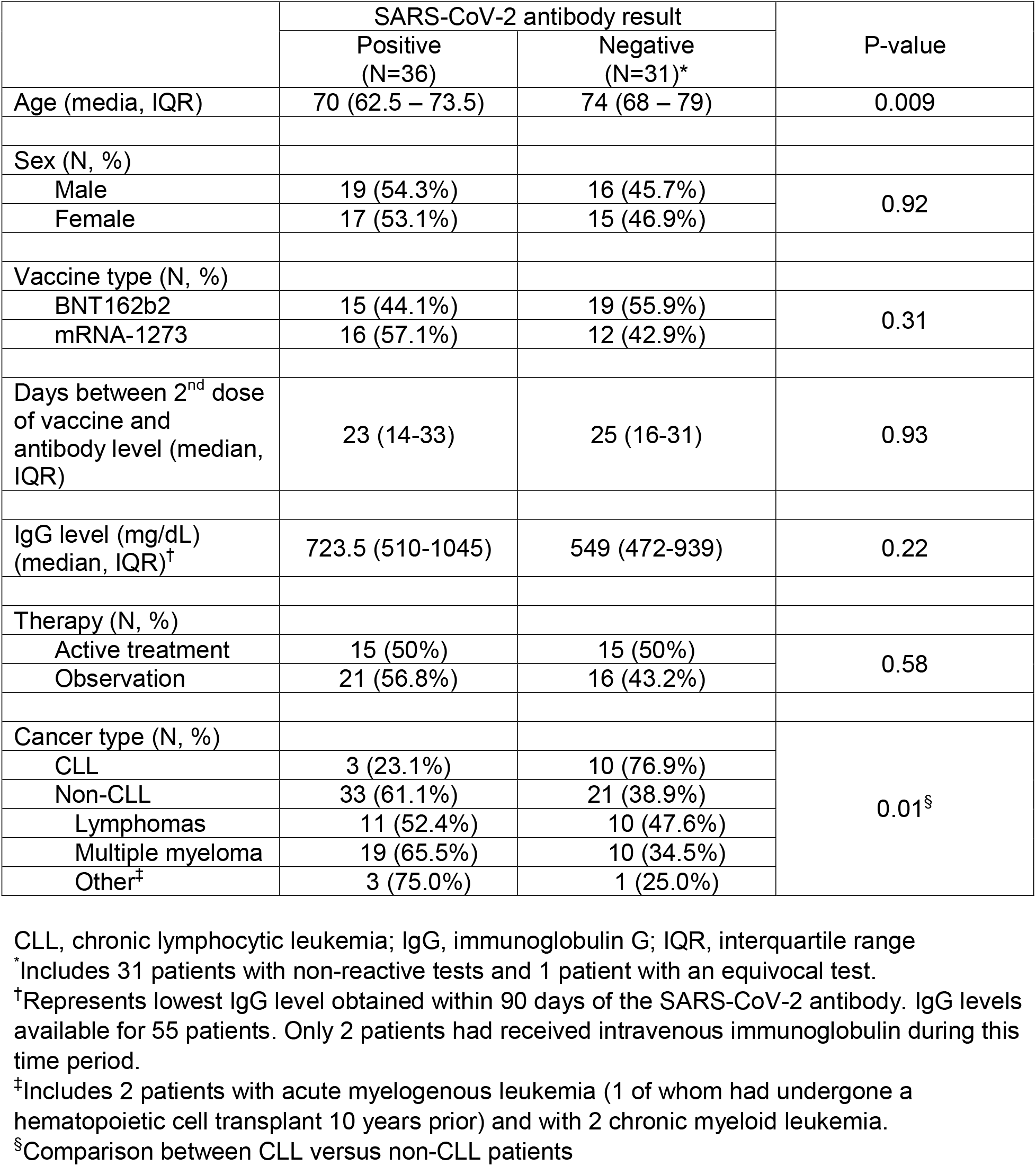
Comparison of hematological malignancy patients with positive versus negative SARS-CoV-2 antibody results after administration of two doses of an mRNA COVID-19 vaccine.

In total, 31/67 patients (46.3%, 95% CI 35.4%– 60.3%) had a negative antibody result after vaccination and were therefore considered to be vaccine non-responders. Older patients were more likely to be vaccine non-responders than younger patients (**Table 1**). Sex, immunoglobulin G (IgG) levels, number of days between 2nd vaccine dose and antibody measurement, and cancer therapy status did not differ among vaccine responders versus non-responders. However, patients with CLL were significantly less likely to develop SARS-CoV-2 antibodies compared to patients with other hematological malignancies (23.1% (3/13) versus 61.1% (33/54), respectively, p = 0.01), even though 69.2% (9/13) of CLL patients were not actively undergoing cancer therapy. There was no difference between age or IgG level between CLL and non-CLL patients.

We further analyzed SARS-CoV-2 IgG extinction coefficient (signal/cutoff) ratios in order to quantify antibody responses. These ratios were obtained from the Beckman assay, with higher values generally indicating more robust antibody responses. Among vaccine responders, there was no difference in the extinction coefficient ratios between the different hematological malignancies (median ratio among CLL versus non-CLL patients = 7.88 (range 1.42 – 20.19) versus 15.44 (range 1.05 – 38.6), respectively, p = 0.39) (**Figure 1A**). Among vaccine non-responders however, patients with CLL had significantly lower extinction coefficient ratios compared to those without CLL (median ratio 0.02 (range 0.02 – 0.06) versus 0.15 (range 0.02 – 0.91), respectively, p < 0.001) (**Figure 1B**). It should be noted that values below 0.10 are suggestive of no antibody response, whereas values closer 1.00 may suggest evolving or declining responses^12^.

**Figure 1.**
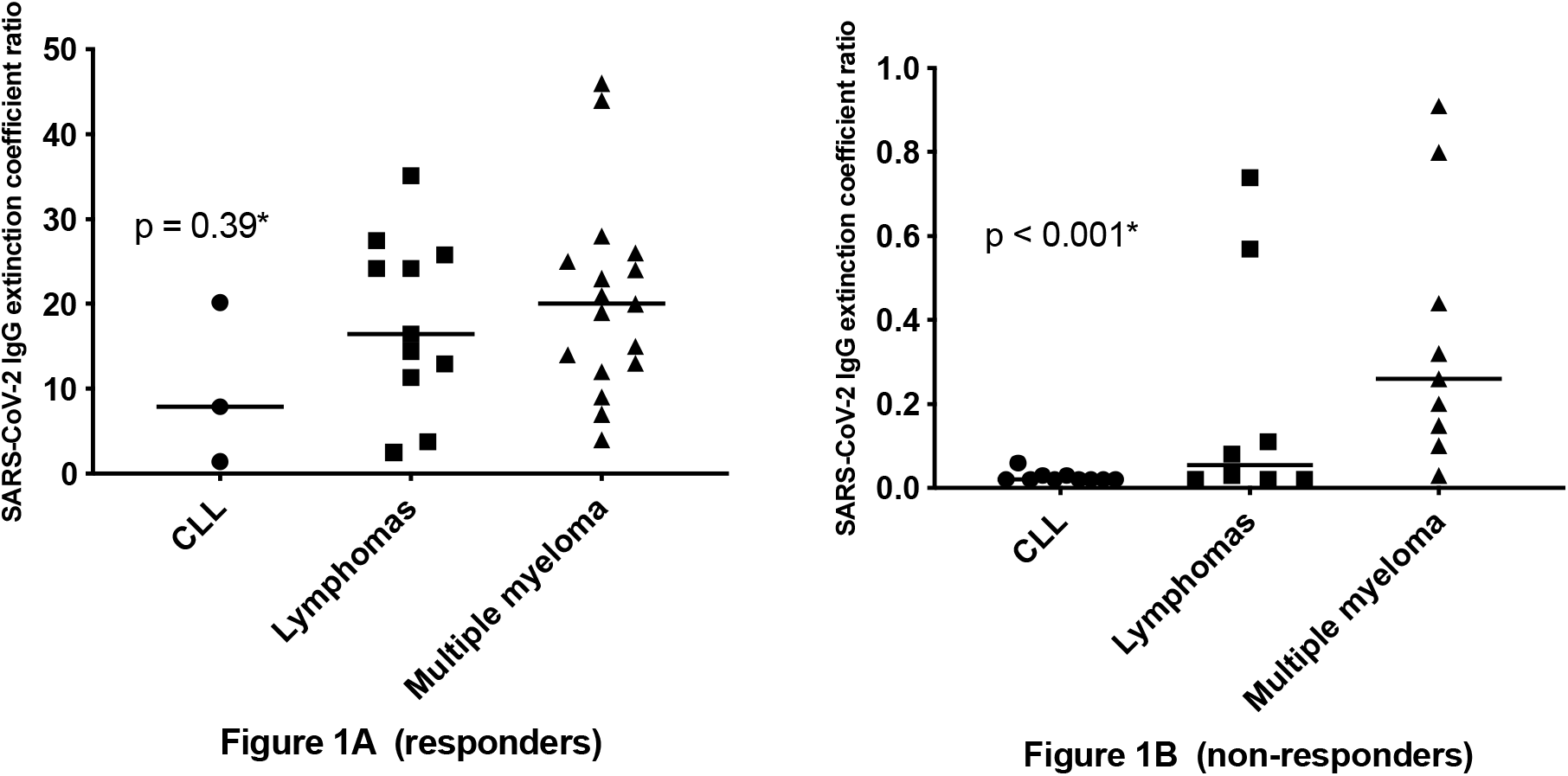
Extinction coefficient (signal/cutoff) ratios of SARS-CoV-2 Spike IgG stratified by vaccine responders (1A) and non-responders (1B), based on whether they had chronic lymphocytic leukemia (CLL) or other hematological malignancies. ^*^Comparisons between CLL versus non-CLL patients. Extinction coefficient ratios for the 4 patients with other myeloid malignancies were not available. Solid lines indicate medians.

Our data show that nearly half of patients with hematological malignancies do not generate antibodies after completing their COVID-19 vaccine series, which is in stark contrast with the results of phase 1 mRNA vaccine immunogenicity trials, in which robust antibody responses were seen in essentially 100% of participants^13,14^. This lack of response was particularly pronounced among patients with CLL, in whom the results of qualitative testing demonstrated significantly lower antibody signals compared to patients without CLL, suggesting that patients with CLL are unable to develop any antibody response after COVID-19 vaccination. These findings cannot be explained by age, cancer therapy, or IgG levels, and are therefore likely a result of the humoral defects that are characteristic of CLL^15^.

Our findings underscore the importance of adherence to non-pharmaceutical interventions to prevent COVID-19 in hematological malignancy patients, particularly in the context of the limited arsenal of SARS-CoV-2 antiviral therapies, the high mortality rates of cancer patients with COVID-19^1-3^, and the emerging risk of prolonged SARS-CoV-2 replication and variant generation in cancer patients^4-7^. Indeed, as the March 2021 CDC guidance has been modified to allow for unmasked gatherings between vaccinated individuals and low-risk unvaccinated individuals^16^, clinicians caring for patients with hematological malignancies and other immunocompromising conditions should be aware of the possibility of COVID-19 vaccine failure. Although immunological correlates of vaccine protection may be more complex than the presence or absence of antibody responses^17^, these patients should be advised to wear masks and observe social distancing regardless of vaccination status.

Limitations of this study include a small sample size, lack of serial measurements, and lack of a control group. In addition, we did not determine whether antibodies from vaccine responders are able to neutralize SARS-CoV-2. Nonetheless, these early findings suggest that COVID-19 vaccine responses in hematological malignancy patient are suboptimal, and that patients with CLL are at a very high risk for vaccine failure. Future studies should focus on post-vaccine antibody durability, B-cell and T-cell responses after vaccination, and novel strategies of COVID-19 prevention in hematological malignancy patients, such as administration of additional vaccine doses or the use of monoclonal antibodies for primary prophylaxis^18^. Routine measurement of SARS-CoV-2 antibody responses in immunocompromised patients should be considered.

## Data Availability

Data are will the first and senior authors and are available upon request.

## Contribution

M.A. and G.H. designed the research and wrote the first draft of the paper. M.A. and M.B. collected the data. G.H. analyzed the data. A.W. and C.C. performed the experiments. All authors have reviewed the paper.

## References

1. Sharma A, Bhatt NS, St Martin A, et al. Clinical characteristics and outcomes of COVID-19 in haematopoietic stem-cell transplantation recipients: an observational cohort study. Lancet Haematol. 2021.

2. Vijenthira A, Gong IY, Fox TA, et al. Outcomes of patients with hematologic malignancies and COVID-19: a systematic review and meta-analysis of 3377 patients. Blood. 2020;136(25):2881–2892.

3. Fung M, Babik JM. COVID-19 in Immunocompromised Hosts: What We Know So Far. Clin Infect Dis. 2021;72(2):340–350.

4. Avanzato VA, Matson MJ, Seifert SN, et al. Case Study: Prolonged Infectious SARS-CoV-2 Shedding from an Asymptomatic Immunocompromised Individual with Cancer. Cell. 2020;183(7):1901–1912 e1909.

5. Aydillo T, Gonzalez-Reiche AS, Aslam S, et al. Shedding of Viable SARS-CoV-2 after Immunosuppressive Therapy for Cancer. N Engl J Med. 2020;383(26):2586–2588.

6. Hensley MK, Bain WG, Jacobs J, et al. Intractable COVID-19 and Prolonged SARS-CoV-2 Replication in a CAR-T-cell Therapy Recipient: A Case Study. Clin Infect Dis. 2021.

7. Kemp SA, Collier DA, Datir RP, et al. SARS-CoV-2 evolution during treatment of chronic infection. Nature. 2021.

8. Ribas A, Sengupta R, Locke T, et al. Priority COVID-19 Vaccination for Patients with Cancer while Vaccine Supply Is Limited. Cancer Discov. 2021;11(2):233–236.

9. Baden LR, El Sahly HM, Essink B, et al. Efficacy and Safety of the mRNA-1273 SARS-CoV-2 Vaccine. N Engl J Med. 2021;384(5):403–416.

10. Polack FP, Thomas SJ, Kitchin N, et al. Safety and Efficacy of the BNT162b2 mRNA Covid-19 Vaccine. N Engl J Med. 2020;383(27):2603–2615.

11. Beckman Coulter Access Immunoassay Systems Instructions for Use. FDA Emergency Use Authorization.. https://www.fda.gov/media/139627/download. Accessed April 2, 2021.

12. Zilla M, Wheeler BJ, Keetch C, et al. Variable Performance in 6 Commercial SARS-CoV-2 Antibody Assays May Affect Convalescent Plasma and Seroprevalence Screening. Am J Clin Pathol. 2021;155(3):343–353.

13. Jackson LA, Anderson EJ, Rouphael NG, et al. An mRNA Vaccine against SARS-CoV-2 -Preliminary Report. N Engl J Med. 2020;383(20):1920–1931.

14. Mulligan MJ, Lyke KE, Kitchin N, et al. Phase I/II study of COVID-19 RNA vaccine BNT162b1 in adults. Nature. 2020;586(7830):589–593.

15. Forconi F, Moss P. Perturbation of the normal immune system in patients with CLL. Blood. 2015;126(5):573–581.

16. When You’ve Been Fully Vaccinated. Centers for Disease Control and Prevention. https://www.cdc.gov/coronavirus/2019-ncov/vaccines/fully-vaccinated.html. Accessed March 26, 2021.

17. Jin P, Li J, Pan H, Wu Y, Zhu F. Immunological surrogate endpoints of COVID-2019 vaccines: the evidence we have versus the evidence we need. Signal Transduct Target Ther. 2021;6(1):48.

18. Lilly’s neutralizing antibody bamlanivimab (LY-CoV555) prevented COVID-19 at nursing homes in the BLAZE-2 trial, reducing risk by up to 80 percent for residents. https://investor.lilly.com/news-releases/news-release-details/lillys-neutralizing-antibody-bamlanivimab-ly-cov555-prevented. Accessed February 18, 2021.

